# COVID-19 Vaccine Hesitancy and Confidence in the Philippines and Malaysia: A Cross-sectional Study of Sociodemographic Factors and Information-Seeking

**DOI:** 10.1101/2022.06.15.22276457

**Authors:** K Brackstone, RR Marzo, R Bahari, MG Head, ME Patalinghug, TT Su

## Abstract

With the emergence of the highly transmissible Omicron variant, large-scale vaccination coverage is crucial to the national and global pandemic response, especially in populous Southeast Asian countries such as the Philippines and Malaysia. Little is known, however, about predictors of COVID-19 vaccine hesitancy and vaccine confidence among unvaccinated individuals in these regions. An internet-based cross-sectional survey was conducted from May 2021 to September 2021. Data from a total of 2558 participants from the Philippines (*N* = 1002) and Malaysia (*N* = 1556) were analysed. Results showed that Filipino (vs. Malaysian) participants indicated higher prevalence of COVID-19 vaccine hesitancy (56.6 vs. 22.9%, *p* = 0.001). However, there were no significant differences in ratings of vaccine confidence between Filipino (45.9%) and Malaysian (49.2%) participants (*p* = 0.105). Predictors associated with greater vaccine hesitancy included females (*p* = 0.029) and rural dwellers (*p* = 0.015) among Filipino participants, whereas females (*p* = 0.004), 25-34 year olds (*p* = 0.027), Christians (*p* < 0.001), and social media use (*p* < 0.001) were associated with hesitancy among Malaysian participants. Predictors associated with lower confidence included females (*p* = 0.026) and information seeking (*p* < 0.001) among Filipino participants, whereas predictors associated with lower confidence among Malaysian participants included residing in a rural community (*p* = 0.004), Christians (*p* < 0.001), online information seeking (*p* < 0.001), and determining relevance of online information (*p* = 0.013). Efforts to improve uptake of COVID-19 vaccination must be centred upon targeting specific communities using local authorities and for the masses through social media. Efforts should focus on determining effective interventions to decrease vaccination hesitancy and increase the uptake of COVID-19 vaccination, particularly in light of the Dengvaxia crisis in the Philippines.

## Introduction

While many high-income settings have achieved relatively high coverage with their COVID-19 vaccination campaigns, almost 35% of the world’s population are still yet to receive a single dose of any COVID-19 vaccine as of May 2022 [1]. The Philippines and Malaysia are among two of the most populous countries in Southeast Asia with an estimated population of 110 million and 32 million people, respectively. Malaysia is doing considerably well with their vaccination efforts, with over 83.3% of the population currently considered fully vaccinated. However, vaccination campaigns in the Philippines have been difficult, with 62.5% of the population currently fully vaccinated [2]. With the emergence of the highly transmissible Omicron variant across the world [3], large-scale vaccination coverage remains fundamental to the national and global pandemic response. Regular scientific assessments of factors that may impede the success of COVID-19 vaccination coverage will be critical as vaccination campaigns continue in these nations.

A key factor for the success of vaccination campaigns is people’s willingness to be vaccinated once doses become accessible to them personally. Vaccine hesitancy is defined by the World Health Organization (WHO) as the delay in the acceptance, or blunt refusal of, vaccines. Hesitancy was described by the WHO as one of the top 10 threats to global health in 2019 [4]. Thus, developing a deeper understanding of the factors associated with vaccine hesitancy and confidence will be crucial toward informing locally-tailored health promotion strategies. Previously, vaccination in Southeast Asia has been associated with mistrust and fear – particularly in the Philippines, who are still suffering the consequences of the Dengvaxia (dengue) vaccine controversy in 2017 [5]. This highly political mainstream event, in which anti-vaccination campaigns linked dengue vaccines with autism spectrum disorder and with corrupt schemes of pharmaceutical companies, may continue to erode the population’s trust in vaccines. For example, a survey conducted on over 30,000 Filipinos in early 2021 showed that 41% of respondents reported that they would refuse the COVID-19 vaccine when it becomes available, whereas Malaysia reported 27% hesitancy [6].

While experts predict that the controversy surrounding Dengvaxia may have weakened the public’s confidence in vaccines [5, 6], there may be many additional factors that also contain negative effects on confidence. The main aims of this research were to determine hesitancy and confidence levels in the COVID-19 vaccine among unvaccinated individuals in the Philippines and Malaysia, and to identify their influencing predictors. This research will help to support the development of locally-tailored health promotion strategies in these regions – especially in light of the Dengvaxia crisis.

## Methods

### Design, subjects, and procedure

This was an internet-based cross-sectional survey conducted from May 2021 to September 2021 in the Philippines and Malaysia. Snowball sampling methods were used for the data collection using social media, including research networks of universities, hospitals, friends, and relatives. Filipino and Malaysian residents aged 18 years old or older were invited to take part. All invited participants consented to the online survey before completion. Consented participants could only respond to questions once using a single account. The voluntary survey contained a series of questions which assessed sociodemographic variables, social media use, digital literacy skills in health, and attitudes toward the COVID-19 vaccine.

### Ethical approval

The study received ethical approval from Asia Metropolitan University’s Medical Research and Ethics Committee (Ref: AMU/FOM/MREC 0320210018). All participants provided informed consent. All study information was written and provided on the first page of the online questionnaire, and respondents indicated consent by selecting the agreement box and proceeding to the survey.

### Measures

#### Demographics

The contents of the questionnaire included socio-demographics such as age, gender, community type (rural vs. urban), educational level (tertiary vs secondary or less), employment (unemployed vs. employed to some degree), religion (Christian, Buddhism, Muslim, Hinduism), income (sufficient vs. insufficient), their perceived ranking on the MacArthur Scale of Subjective Social Status [7], whether they had a chronic disease (no vs. yes), whether they were permanently impaired by a health problem (no vs. yes), and whether they were social media users (no vs. yes).

#### Vaccine confidence and hesitancy

Participants were also asked about their perceived level of confidence in the COVID-19 vaccine (“I am completely confident that the COVID-19 vaccine is safe,” 1 = *strongly disagree*; 7 = *strongly agree*). Then, participants were asked about their level of hesitancy to the COVID-19 vaccine (“I think everyone should be vaccinated according to the national vaccination schedule”; no, I don’t know, yes).

#### Digital Health Literacy

Finally, participants completed the Digital Health Literacy Instrument (DHLI) [8], which was adapted in the context of the COVID-HL Network. The scale measures one’s ability to seek, find, understand, and appraise health information from digital resources. While the original DHLI is comprised of 7 subscales, we used the following four domains including: (1) information searching or using appropriate strategies to look for information (e.g., “When you search the internet for information on coronavirus virus or related topics, how easy or difficult is it for you to find the exact information you are looking for?”) (2) adding self-generated content to online-based platforms (e.g., “When typing a message on a forum or social media such as Facebook or Twitter about the coronavirus a related topic, how easy or difficult is it for you to express your opinion, thought, or feelings in writing?”) (3) evaluating reliability of online information (e.g., “When you search the internet for information on the coronavirus or related topics, how easy or difficult is it for you to decide whether the information is reliable or not?”) and (4) determining relevance of online information (e.g., “ When you search the internet for information on the coronavirus or related topics, how easy or difficult is it for you to use the information you found to make decisions about your health (protective measures, hygiene regulations, transmission routes, risks and their prevention?”). A total of 12 items (three per each dimension) were asked, and answers were recorded on a four-point Likert scale (1 = *very difficult*; 4 = *very easy*).

### Data analysis

Data were examined for errors, cleaned, and exported into IBM SPSS Statistics 28 for further analysis. All hypotheses were tested at a significance level of 0.05. *χ*^2^ tests were conducted for group differences of categorical variables, and Mann-Whitney tests for continuous variables. Subgroup analyses were performed for Filipino and Malaysian participants.

COVID-19 vaccine hesitancy and confidence were treated as separate dependent variables in a logistic regression model providing the strictest test of potential influence of COVID-19 vaccine hesitancy and confidence among Filipino and Malaysian participants. Independent variables were: age, gender, community type, educational level, employment, religion, income, perceived ranking on the MacArthur Scale of Subjective Social Status [7], chronic disease presence, health impairment presence, social media use, and the four domains of the DHLI scale [8]. Low vaccine confidence was operationalised by dichotomising participants’ responses to the statement: “I am completely confident that the COVID-19 vaccine is safe” into those who disagreed or neither agreed nor disagreed (1-4) vs. high vaccine confidence (5-7). Vaccine hesitancy was operationalised by dichotomising responses to the statement: “I think everyone should be vaccinated according to the National vaccination schedule” into those indicating ‘no’ or ‘I don’t know’ (vs. ‘yes’ indicating low vaccine hesitancy).

## Results

A total of 2558 participants completed the online survey. Table 1 shows descriptive statistics of participants from the Philippines (*N* = 1002) vs. Malaysia (*N* = 1556). Filipino (vs. Malaysian) participants indicated higher rates of education (*p* < 0.001), but were more likely to be unemployed (*p* < 0.001). Further, Filipino (vs. Malaysian) participants were also more likely to indicate insufficient income (*p* < 0.001) and rate themselves lower on the socioeconomic scale (*p* < 0.001). Malaysian (vs. Filipino) participants were more likely to live in urban areas (*p* < 0.001). Most notably, Filipino participants indicated higher prevalence of COVID-19 vaccine hesitancy compared to Malaysian participants (56.6 vs. 22.9%, *p* = 0.001). However, there were no significant differences between Filipino (45.9%) and Malaysian (49.2%) participants in ratings of vaccine confidence (*p* = 0.105). Malaysian (vs. Filipino) participants were also more likely to report using social media (96.6 vs. 89.8%; < 0.001).

**Table 1.** Socio-demographic characteristics of participants from the Philippines vs. Malaysia. Values are presented as percent (n) or means ± SD.

| Characteristics | Total (N = 2558) | Philippines (N = 1002) | Malaysia (N = 1556) | *p* |
| --- | --- | --- | --- | --- |
| **Gender** |  |  |  | < 0.001 |
| Female | 54.1 (1384) | 60.8 (608) | 49.8 (775) |  |
| Male | 45.9 (1174) | 39.2 (393) | 50.2 (581) |  |
| **Age** |  |  |  | < 0.001 |
| 18-24 | 72.2 (1846) | 66.3 (845) | 64.3 (1001) |  |
| 25-34 | 19.1 (488) | 12.2 (122) | 23.5 (266) |  |
| 35-44 | 8.7 (224) | 3.5 (35) | 12.2 (189) |  |
| **Community type** |  |  |  | < 0.001 |
| Rural | 37.5 (958) | 66.3 (664) | 18.9 (294) |  |
| Urban | 62.5 (1600) | 33.7 (338) | 81.1 (1262) |  |
| **Highest level of education** |  |  |  | < 0.001 |
| Senior secondary or lower | 40.2 (1029) | 30.7 (308) | 46.3 (721) |  |
| Tertiary | 59.8 (1529) | 69.3 (394) | 53.7 (835) |  |
| **Employment** |  |  |  | < 0.001 |
| Unemployed | 72.9 (1864) | 81.4 (826) | 67.4 (1048) |  |
| Employed or self-employed | 27.1 (694) | 18.6 (186) | 32.6 (508) |  |
| **Religion** |  |  |  | < 0.001 |
| Islam | 35.3 (904) | 2.4 (24) | 57.0 (880) |  |
| Christian | 43.3 (1108) | 92.7 (929) | 11.6 (179) |  |
| Buddhism | 9.1 (233) | 0.1 (1) | 15.0 (232) |  |
| Hinduism | 11.7 (300) | 4.8 (48) | 16.3 (252) |  |
| **Income** |  |  |  | < 0.001 |
| Sufficient | 72.9 (1865) | 62.3 (624) | 79.8 (1241) |  |
| Insufficient | 27.1 (693) | 37.7 (378) | 20.2 (315) |  |
| **Socioeconomic scale** | 6.23 ± 1.85 | 5.89 ± 1.92 | 6.45 ± 1.78 | < 0.001 |
| **Chronic disease** |  |  |  | < 0.001 |
| No | 92.3 (2392) | 89.7 (899) | 94.0 (1463) |  |
| Yes | 7.7 (196) | 10.3 (103) | 6.0 (93) |  |
| **Health impairment** |  |  |  | 0.055 |
| No | 85.5 (2186) | 80.6 (808) | 88.6 (1378) |  |
| Yes | 14.5 (372) | 19.4 (194) | 11.4 (178) |  |
| **Vaccine hesitancy** |  |  |  | < 0.001 |
| Low | 63.9 (1634) | 43.4 (435) | 77.1 (1199) |  |
| High | 36.2 (954) | 56.6 (567) | 22.9 (357) |  |
| **Vaccine confidence** |  |  |  | < 0.105 |
| Low | 52.1 (1332) | 54.1 (542) | 50.8 (790) |  |
| High | 47.9 (1226) | 45.9 (460) | 49.2 (766) |  |
| **Social media use** |  |  |  | < 0.001 |
| No | 6.04 (155) | 10.2 (102) | 3.4 (53) |  |
| Yes | 93.6 (2403) | 89.8 (900) | 96.6 (1503) |  |
| **Digital Health Literacy** |  |  |  |  |
| Information seeking | 2.15 ± 0.65 | 2.29 ± 0.57 | 2.07 ± 0.68 | < 0.001 |
| Self-generated content | 2.15 ± 0.65 | 2.29 ± 0.55 | 2.07 ± 0.69 | < 0.001 |
| Evaluating reliability | 2.20 ± 0.70 | 2.37 ± 0.57 | 2.11 ± 0.74 | < 0.001 |
| Determining relevance | 2.09 ± 0.68 | 2.22 ± 0.57 | 2.02 ± 0.73 | < 0.001 |

Table 2 shows significant predictors of vaccine hesitancy in both Filipino and Malaysian samples. Among Filipino participants, multivariate logistic regression analyses revealed that factors associated with higher vaccine hesitancy included females (OR, 1.51, 95% CI, 1.14-2.00; *p* = 0.004), residing in a rural community (OR, 1.45, 95% CI, 1.07-1.95; *p* = 0.015), and having insufficient income (OR, 1.62, 95% CI, 1.20-2.19; *p* = 0.001). Among Malaysian participants, females (OR, 1.51, 95% CI, 1.14-2.00; *p* = 0.004), being aged 25-34 (vs. 18-24; OR, 1.52, 95% CI, 1.48-2.21; *p* = 0.027), Christians (OR, 2.45, 95% CI, 1.66-3.62; *p* < 0.001), completing tertiary education (OR, 2.17, 95% CI, 1.63-2.88; *p* < 0.001), social media use (OR, 11.59, 95% CI, 5.63-23.84; *p* < 0.001), and information seeking (OR, 2.50, 95% CI, 1.74-3.61; *p <* 0.001) were predictors of higher vaccine hesitancy, whereas factors associated with lower vaccine hesitancy included having a health impairment (OR, 0.49, 95% CI, 0.30-0.78; *p* = 0.003) and higher self-reported ratings on the socioeconomic scale (OR, 0.82, 95% CI, 0.75-0.89; *p* < 0.001).

**Table 2.** Multivariate conditional logistic regression analysis of vaccine hesitancy in the Philippines and Malaysia

| Variables | OR | 95% CI | p-value |
| --- | --- | --- | --- |
| **The Philippines** |  |  |  |
| Female | 1.378 | 1.034 – 1.838 | .029 |
| Rural community | 1.451 | 1.075 – 1.959 | .015 |
| Insufficient income | 1.625 | 1.205 – 2.192 | .001 |
| **R<sup>2</sup>** |  |  | **0.062** |
| **Malaysia** |  |  |  |
| Female | 1.514 | 1.145 – 2.003 | 0.004 |
| 25-34 | 1.523 | 1.048 – 2.214 | 0.027 |
| Christian beliefs | 2.458 | 1.664 – 3.629 | <0.001 |
| Higher education (tertiary) | 2.173 | 1.634 – 2.889 | <0.001 |
| Health impairment | 0.490 | 0.306 – 0.786 | 0.003 |
| Socioeconomic scale | 0.824 | 0.755 – 0.899 | <0.001 |
| Social media use | 11.591 | 5.634 – 23.845 | <0.001 |
| Information seeking | 2.507 | 1.741 – 3.610 | <0.001 |
| **R<sup>2</sup>** |  |  | **0.281** |
| **Total** |  |  |  |
| Female | 1.428 | 1.174 – 1.736 | < 0.001 |
| Urban community | 1.531 | 1.240 – 1.889 | < 0.001 |
| Christian beliefs | 3.526 | 2.805 – 4.433 | < 0.001 |
| Hindu | 0.644 | 0.416 – 0.996 | 0.048 |
| Higher education | 1.530 | 1.254 – 1.868 | <0.001 |
| Insufficient income | 1.423 | 1.147 – 1.766 | 0.001 |
| Health impairment | 0.691 | 0.519 – 0.919 | 0.011 |
| Social media use | 6.987 | 3.848 – 12.688 | <0.001 |
| Socioeconomic scale | 0.893 | 0.845 – 0.843 | <0.001 |
| Information seeking | 1.739 | 1.340 – 2.257 | <0.001 |
| **R<sup>2</sup>** |  |  | **0.271** |

*OR, odds ratio; CI, confidence interval. Multivariate logistic regression analyses using stepwise variable selection.*

Table 3 shows significant predictors of vaccine confidence in both Filipino and Malaysian samples. Factors positively associated with higher vaccine confidence among Filipino participants included higher self-reported ratings on the socioeconomic scale (OR, 1.16, 95% CI, 1.07-1.25; *p* < 0.001), whereas factors associated with lower vaccine confidence included females (OR, 0.72, 95% CI, 0.54-0.96; *p* = 0.026) and information seeking (OR, 0.63, 95% CI, 0.49-0.81; *p* < 0.001). Among Malaysian participants, factors positively associated with higher vaccine confidence included females (OR, 1.27, 95% CI, 1.18-1.60; *p* = 0.035), completing tertiary education (OR, 1.31, 95% CI, 1.03-1.66; *p* = 0.026), and higher self-reported ratings on the socioeconomic scale (OR, 1.08, 95% CI, 1.00-1.16; *p* = 0.036). Factors negatively associated with lower vaccine confidence included residing in a rural community (OR, 0.63, 95% CI, 0.47-0.87; *p* = 0.004), Christians (OR, 0.50, 95% CI, 1.20-2.24; *p* < 0.001), Buddhists (OR, 0.15., 95% CI, 0.10-0.22; *p* < 0.001), Hindus (OR, 0.24., 95% CI, 0.17-0.34; *p* = 0.004), information seeking (OR, 0.42, 95% CI, 0.31-0.58; *p* < 0.001), and determining relevance of online information (OR, 0.68, 95% CI, 0.51-0.92; *p* = 0.013).

**Table 3.**
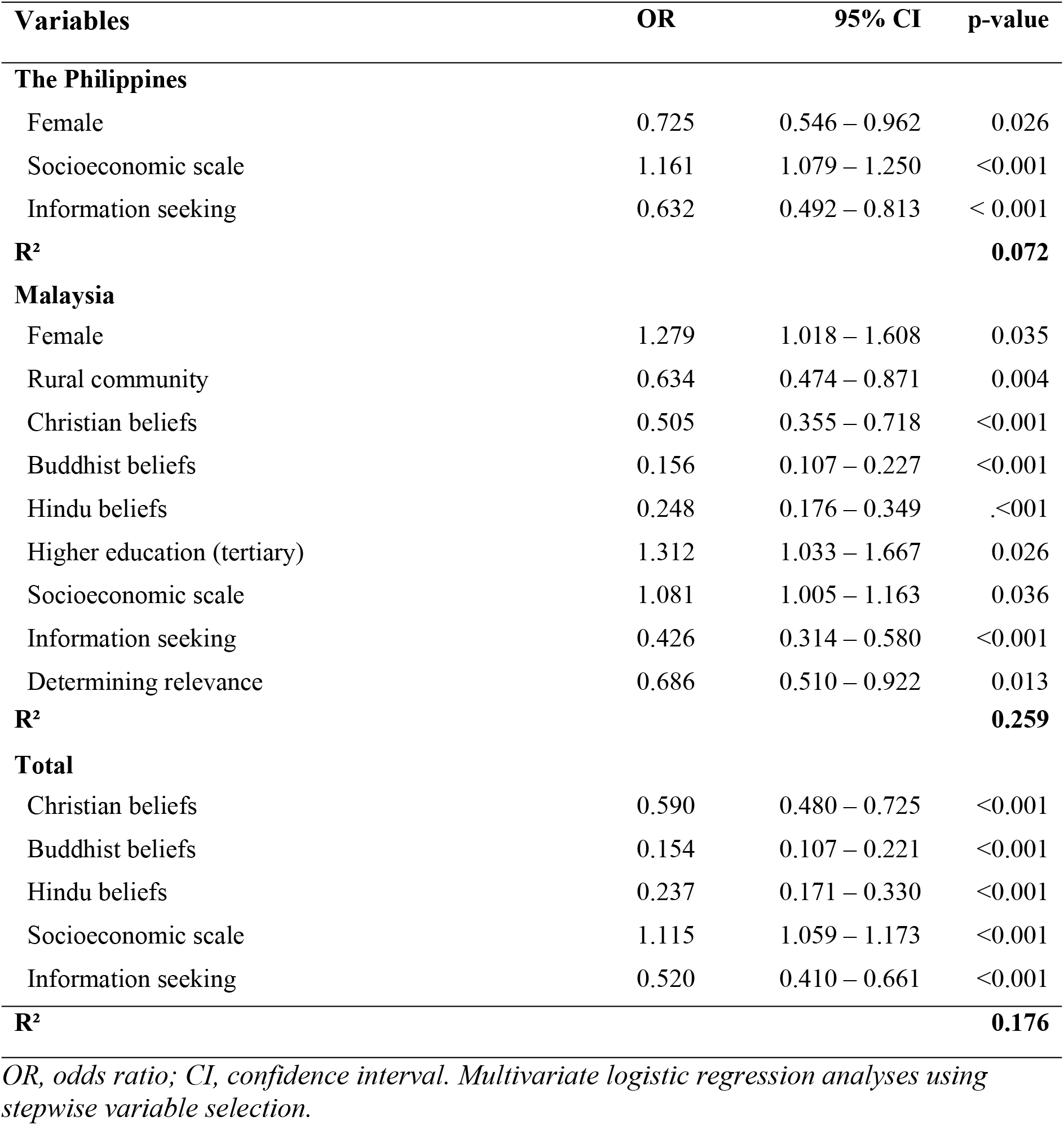
Multivariate conditional logistic regression analysis of vaccine confidence in the Philippines and Malaysia – significant associations

| Variables | OR | 95% CI | p-value |
| --- | --- | --- | --- |
| **The Philippines** |  |  |  |
| Female | 0.725 | 0.546 – 0.962 | 0.026 |
| Socioeconomic scale | 1.161 | 1.079 – 1.250 | <0.001 |
| Information seeking | 0.632 | 0.492 – 0.813 | < 0.001 |
| **R<sup>2</sup>** |  |  | **0.072** |
| **Malaysia** |  |  |  |
| Female | 1.279 | 1.018 – 1.608 | 0.035 |
| Rural community | 0.634 | 0.474 – 0.871 | 0.004 |
| Christian beliefs | 0.505 | 0.355 – 0.718 | <0.001 |
| Buddhist beliefs | 0.156 | 0.107 – 0.227 | <0.001 |
| Hindu beliefs | 0.248 | 0.176 – 0.349 | .<001 |
| Higher education (tertiary) | 1.312 | 1.033 – 1.667 | 0.026 |
| Socioeconomic scale | 1.081 | 1.005 – 1.163 | 0.036 |
| Information seeking | 0.426 | 0.314 – 0.580 | <0.001 |
| Determining relevance | 0.686 | 0.510 – 0.922 | 0.013 |
| **R<sup>2</sup>** |  |  | **0.259** |
| **Total** |  |  |  |
| Christian beliefs | 0.590 | 0.480 – 0.725 | <0.001 |
| Buddhist beliefs | 0.154 | 0.107 – 0.221 | <0.001 |
| Hindu beliefs | 0.237 | 0.171 – 0.330 | <0.001 |
| Socioeconomic scale | 1.115 | 1.059 – 1.173 | <0.001 |
| Information seeking | 0.520 | 0.410 – 0.661 | <0.001 |
| **R<sup>2</sup>** |  |  | **0.176** |

*OR, odds ratio; CI, confidence interval. Multivariate logistic regression analyses using stepwise variable selection.*

## Discussion

Malaysia and the Philippines are among the top emerging economies in Southeast Asia. Both countries are also among the most populous in the region. However, while the economic impact of the COVID-19 pandemic has been permanent in the Philippines, it has been shown thus far to be temporary in Malaysia [9]. Between January and October 2020, about 30,000 Malaysians had been infected by the virus with a mortality rate of 0.79%, while approximately 380,000 cases of COVID-19 were detected in the Philippines with a mortality rate of 1.9% [10]. Further, 61.8% of Malaysians had completed their vaccination up until September 2021, while the percentage of completed vaccinations during the same period in the Philippines was only 19.2% [11]. Vaccine uptake is likely to be a key determining factor in the outcome of a pandemic. Knowledge around factors predicting vaccine hesitancy and confidence is of the utmost important in order to improve vaccination rates.

In this study, a number of important findings on vaccine hesitancy and confidence in the Philippines and Malaysia are described. First, while there were no significant differences in ratings of confidence in the COVID-19 vaccine between Filipino and Malaysian participants, Filipino (compared to Malaysian) participants expressed greater vaccine hesitancy. This may be a consequence of previous vaccine scares in the years leading up to the pandemic, including the Dengvaxia controversy in 2016 [5]. Systematic reviews demonstrated that, by the end of 2020, the highest vaccine acceptance was in China, Malaysia, and Indonesia [12, 13]. The authors postulated that this elevated awareness was due to being among the first countries affected by the virus, hence resulting in greater confidence in vaccines [12].

Second, this study shows that being female is a common predictor for vaccine hesitancy in both countries. The evidence base shows mixed findings, with other studies reporting greater hesitancy in females [14, 15] or in males [15]. In some countries, the gender gap is not as substantial as others. In a large global study conducted in countries such as Russia and the United States, it was found that there is greater gender gap in vaccine hesitancy among males and females compared to countries such as Nepal and Sierra Leone [17, 18]. Unsurprisingly, what drives this higher hesitancy in some studies is the inclusion of pregnant females, where it had been consistently demonstrated that this population is more hesitant at taking getting vaccinated due to concerns for their babies [19]. Hence, after taking all consideration into account, gender differences in vaccine hesitancy cannot be supported with certainty. This also emphasises the need for tailored health promotion towards the key populations at risk.

There are clear differences in predictors of vaccine hesitancy in the Philippines and Malaysia. However, when results for both countries were combined, females, urban dwellers, those of Christian faith, those with higher educational attainment, higher self-reported social class, greater use of social media, and information seeking tendencies remained as predictors of hesitancy. Urban-dwellers and individuals with more years of education have previously been demonstrated as predictors for vaccine hesitancy in previous studies [20], but contradictory results have also previously been shown [21, 22]. Nevertheless, reports have shown higher vaccine refusals among those with strong religious beliefs such as the Amish Community in the United States and the Orthodox Protestants in the Netherlands [23], as well as some Muslim groups in Pakistan [24].

Frequent use of social media is the only strong predictor for vaccine hesitancy in this study, followed by information seeking. Research has identified that the safety and effectiveness of the vaccine is the primary concern that people have, including beliefs in available information [25, 26, 27]. Unfortunately, high internet literacy is a double-edged sword in this case, since participants in this study preferred to seek information through social media, and thus be exposed to inaccurate information regarding COVID-19 vaccine. Previous studies have associated higher vaccine hesitancy with misinformation about the virus and vaccines [24], particularly if they relied on social media as a key source of information [28]. A 2022 systematic review discovered that high social media use is the main driver of vaccine hesitancy across all countries around the globe, and is especially prominent in Asia [29]. Furthermore, vaccine acceptance and uptake improved among those who obtained their information from healthcare providers compared to relatives or the internet [30].

In terms of vaccine acceptance, our findings show that those with better perceived socioeconomic standing have higher confidence in vaccination, consistent with previous studies describing how those with a higher income had expressed willingness to pay for their COVID-19 vaccination if necessary [17, 31, 32]. Further, those of Christian, Buddhist, and Hindu faiths, as well as those with a tendency to seek information, were associated with lower vaccine confidence. This is in keeping with the previous findings demonstrating that strong religious convictions are often tied to mistrust of authorities and beliefs about the cause of the COVID-19 pandemic, which is fuelled by social media [33]. Furthermore, concern on the permissibility of these vaccines in their religion reduces its acceptability [34]. However, it is interesting to note that, while the majority in Malaysia are Muslims, it did not reduce the rate of vaccine acceptance and confidence in the country.

These findings are useful to inform health authorities and governments on the areas to focus on to improve vaccination uptake. Misinformation about vaccination greatly hampers vaccination efforts, hence why there is a need to spread awareness by providing the right information driven by governmental and non-governmental organisations [29]. This could be achieved by having community-specific public education and role modelling from local health and public officials, which has been shown to increase public trust [35]. Since the primary reason for hesitancy is concern about the safety of vaccines, it is crucial that education programmes continue to stress the effectiveness and importance of COVID-19 vaccinations [36]. Participants in this study coped with the pandemic by seeking information, but they sought information from social media when information from the authorities was lacking or were viewed as untrustworthy, which may have contained erroneous information. One way to deter this is to empower information-technology companies to monitor vaccine related materials on social media, remove false information, and create correct and responsible contents [35]. Furthermore, behavioural change techniques have been found to be useful in stressing the consequences of rejecting the vaccine on physical and mental health [37]. The most effective “nudging” interventions include offering incentives for parents and healthcare workers, providing salient information, and employing trusted figures to deliver this information [38]. Finally, since religious concerns have been prominent in reducing vaccine confidence and increasing hesitancy in this study, it is important to tailor messages to include information related to religion, and the use of religious leaders to spread these messages [39]. These are all important factors for increasing uptake of the COVID-19 vaccine, but also may be relevant in acceptability of routine immunisations as countries look to transition towards a post-pandemic delivery of healthcare.

A limitation of this study include its cross-sectional design and the heterogeneity among participants. However, the large sample sizes included in this study offsets this limitation and renders support for our conclusions.

## Conclusions

Predictors of vaccine hesitancy in this study include the use of social media, information seeking, and Christianity. Higher socioeconomic status positively predicts vaccine confidence; however, being Christian, Buddhist or Hindu, and the tendency to seek information are predictors of hesitancy. Efforts to improve uptake of COVID-19 vaccination must be centred upon providing accurate information to specific communities using local authorities, health services and other locally-trusted voices (such as religious leaders), and for the masses through social media. Further studies should focus on determining effective interventions to improve vaccination confidence and increase the uptake of vaccination.

## Data Availability

Data are available on the OSF: https://osf.io/ncwjq/

